# Relapse prediction in multiple myeloma patients treated with isatuximab, carfilzomib, and dexamethasone

**DOI:** 10.1101/2024.05.02.24306607

**Authors:** Even Moa Myklebust, Fredrik Schjesvold, Arnoldo Frigessi, Kevin Leder, Jasmine Foo, Alvaro Köhn-Luque

## Abstract

Multiple myeloma (MM) patients experience repeated cycles of treatment response and relapse, yet despite close monitoring of disease status through M protein measurements, no standard model exists for relapse prediction in MM. We investigate the feasibility of predicting relapse using a hierarchical Bayesian model of subpopulation dynamics by training and testing the model on 229 patients from the IKEMA trial.

After observing between 11 and 18 treatment cycles, the model predicted relapse within six cycles with an average sensitivity between 60 and 80 %, and an average specificity between 60 and 90 %. A model of linear extrapolation is preferable when patients have been observed for less than 6 cycles, but for longer observation windows the hierarchical Bayesian model is preferred. Including available baseline and longitudinal covariate information did not improve predictive accuracy. A survival analysis showed that two model parameters separated patients into groups with significantly different PFS (*p <* 0.001).

**Statement of Significance:** Currently, no standard model exists for relapse prediction in multiple myeloma. A personalized model of M protein development could guide the frequency of follow-up measurements, reduce uncertainty for patients, and give clinicians more time to choose the best subsequent treatment for each patient. Furthermore, models that predict relapse are required to study the effect of changing treatment in advance of relapse rather than in response to it. Our work addresses this need by developing a hierarchical Bayesian model of subpopulation dynamics for prediction of future M protein values. We validate the model on a patient cohort treated with state-of-theart CD38 inhibitor therapy and show that it can accurately predict relapse within the next six treatment cycles, highlighting the promise of mathematical modeling in multiple myeloma and for personalized medicine in general.

**Declaration of Interests:** F.S. received honorarium from Sanofi, Janssen, BMS, Oncopeptides, Abbvie, GSK, and Pfizer. The authors declare that they have no other conflicts of interest.

## Introduction

Multiple myeloma (MM) is a hematologic malignancy that affects about 150,000 people annually worldwide [1]. It develops from monoclonal gammopathy of undetermined significance (MGUS), an asymptomatic preneoplastic plasma cell disorder that is characterized by an elevated level of monoclonal antibodies known as M protein. Patients who develop MM typically experience anemia, bone lesions, and potentially hypercalcemia and reduced kidney function [2]. For newly diagnosed MM patients, the standard care is induction therapy with a proteasome inhibitor followed by high-dose melphalan treatment, autologous hematopoietic stem cell transplantation, and lenalidomide maintenance therapy [3]. Novel treatment options for relapsed and refractory patients include CD38-antibodies daratumumab [4] and isatuximab [5], typically combined with a proteasome inhibitor and dexamethasone [6, 7].

Despite the introduction of new treatments, only 10-15 % of patients achieve or exceed the expected survival of the matched general population [3]. A key reason for this is the large intra-tumor heterogeneity in MM [8]. Due to their wide genomic variation, heterogeneous cancers are more likely to harbor resistant subpopulations [9]. Large inter-patient heterogeneity in response time [8] further complicates the situation. Yet apart from the chromosomal translocation t(11;14), which is a predictive biomarker for BCL-2 therapy [10], prognostic factors and molecular markers for personalized treatment are largely lacking in MM. Consequently, there is both a potential and a need to identify molecular markers of relapse risk in MM.

Mathematical models have a long-standing history of guiding treatment decisions and contributing to the understanding of underlying biological mechanisms by quantifying bio-medical assumptions and making them testable [11]. The presence of M protein as a proxy of tumor burden has made MM in particular a target for efforts to model treatment response and search for optimal therapies [12, 13]. More recent work has attempted to predict future M protein trajectories using mathematical modeling [14, 15]. Related work in head-and-neck cancer has used statistical modeling of longitudinal tumor burden measurements to predict tumor volume progression [16]. In this work, we propose a hierarchical Bayesian model of subpopulation dynamics that builds on these ideas by including automatic discovery of covariate effects.

Repeated observations of M protein provide opportunities for statistical inference about the underlying population dynamics. We present a hierarchical Bayesian model of subpopulation dynamics which predicts future M protein values by modeling the decay and growth rates of sensitive and resistant subpopulations, and which is also capable of incorporating information from covariates measured at baseline or during treatment. The model employs a nonlinear mixed effect model (NLME) [17] framework which handles observation noise, allows sharing of information between patients, and uses sparsity-enforcing priors to discover covariate effects without overfitting. The model predicts future M protein values on an individual level through combining observed individual M protein measurements with trends in the wider population.

## Materials and methods

### Study population and exclusions

In this work, we studied a subset of patients from the multicenter, phase III, open-label trial IKEMA [18]. In the trial, 302 patients with relapsed or refractory MM, who had received one to three lines of prior therapy, were randomly assigned (3:2) to treatment with either isatuximab plus carfilzomib and dexamethasone (isatuximab group) or carfilzomib and dexamethasone (control group). In total, 179 patients were assigned to the isatuximab group and 123 patients to the control group. Non-secretory patients were excluded from entry into the trial.

From this population, patients with less than three non-zero M protein measurements were excluded in this work to obtain reliable parameter estimates for every patient. This reduced the population size to 229 patients: 134 in the isatuximab group and 95 in the control group. Baseline covariates and longitudinal M protein measurements for patients in both groups were used in the analysis.

### Mathematical model of treatment response and relapse

In 2008, Stein et. al. [19] presented a method for predicting survival in prostate cancer by estimating decay and growth rates of serum prostate-specific antigen (PSA). The total PSA level was modeled as the sum of PSA contributions from two populations: a decaying population (sensitive to treatment) and a growing population (resistant to treatment). Equation (1) describes their model, with *ρ*^*r*^ as the growth rate of the resistant subpopulation and *ρ*^*s*^ as the (negative) decay rate of the sensitive subpopulation.

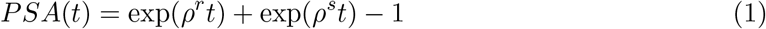

In their study of 112 patients, the log value of the growth rate was found to be strongly correlated with survival. A larger retrospective study [20] also found PSA growth rate to be predictive of overall survival. Subsequent work included a parameter for the proportion of the resistant subpopulation and used the extended model to estimate growth and decay rates from CT volume measurements in colorectal cancer [21]. Equation (2) shows an equivalent version of that model, where *π*_*r*_ is the proportion of resistant cells.

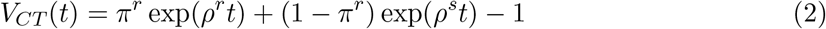

The same rationale can be applied to multiple myeloma. Using a slightly different model, the total M protein at time *t* is described in equation (3), where *M* ^0^ is the initial total amount of M protein, *π*^*r*^ is the initial proportion of resistant cells, *ρ*^*r*^ is the growth rate of the resistant subpopulation, and *ρ*^*s*^ (negative) is the decay rate of the sensitive subpopulation.

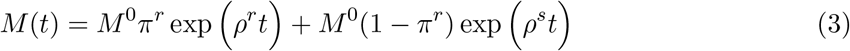

The two populations are assumed to secrete M protein at the same time-constant rate. The assumption that the resistant subpopulation grows exponentially is made plausible by the fact that patients who progress are removed from the study before the carrying capacity is reached. Together, the model parameters *ρ*^*r*^, *ρ*^*s*^, *π*^*r*^, and *M* ^0^ uniquely describe an M protein trajectory. Figure 1 illustrates the role of each of the parameters in the model.

**Figure 1.**
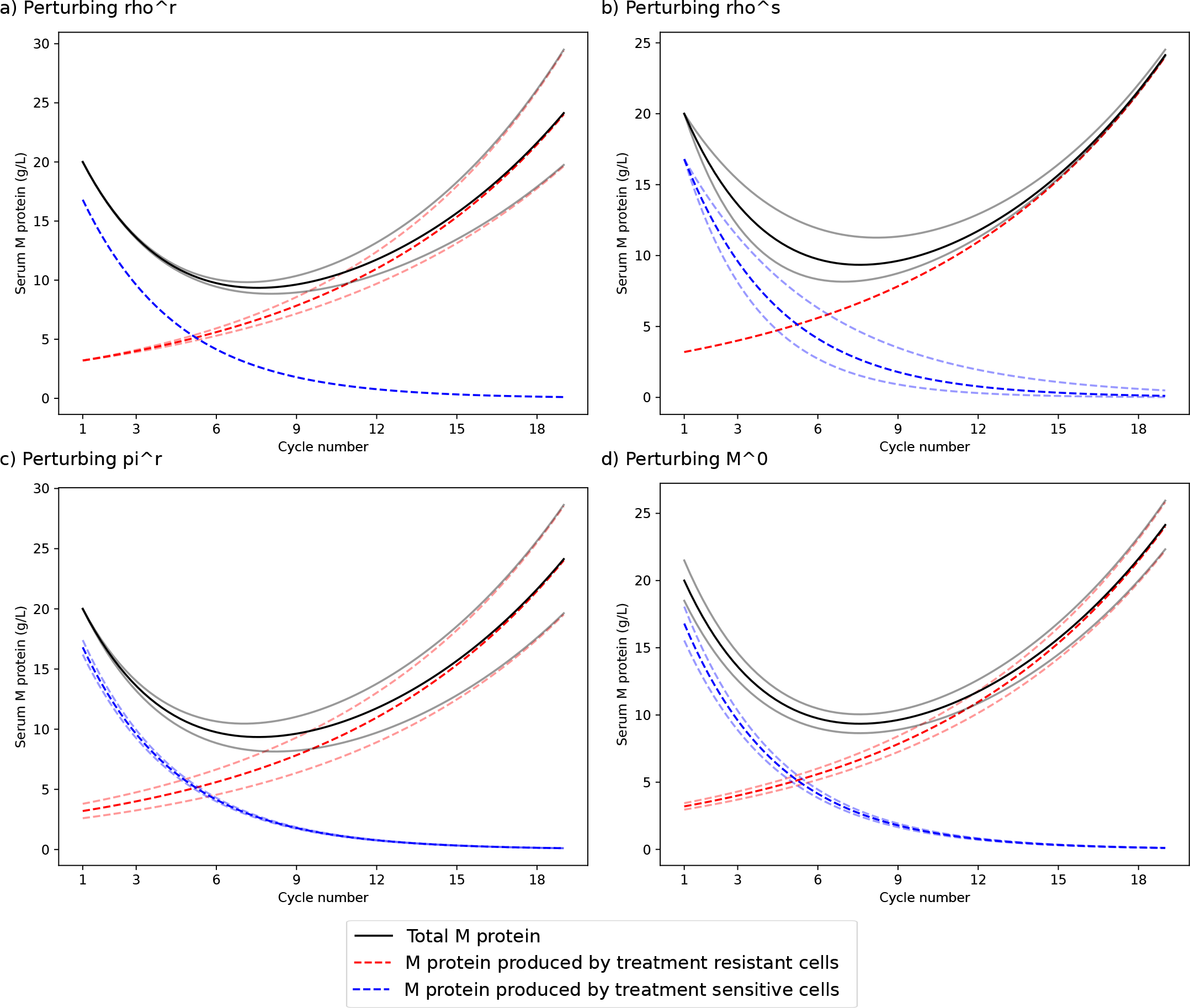
Influence of model parameters on M protein trajectory. This figure shows the influence of small changes in the model parameters in equation (3) on the M protein trajectory of a patient with growth rate of the resistant population *ρ*^*r*^ = 0.004; decay rate of the sensitive population *ρ*^*s*^ = − 0.01; proportion of resistant cells at treatment start *π*^*r*^ = 0.16; and M protein level at treatment start *M* ^0^ = 20. **a)** *ρ*^*r*^ is changed from 0.004 to 0.0036 and 0.0044, respectively. **b)** *ρ*^*s*^ is changed from *−*0.01 to *−*0.007 and *−*0.013, respectively. **c**) *π*^*r*^ is changed from 0.16 to 0.11 and 0.21, respectively. **d)** *M*^0^ is changed from 20 to 18.5 and 21.5, respectively.

### Covariate effects

For a single patient, the model parameters *ρ*^*r*^, *ρ*^*s*^, *π*^*r*^, and *M* ^0^ can be estimated from observed M protein values. As M protein observations are typically sparse in time and come with measurement errors, some level of uncertainty in each parameter estimate is to be expected. It can therefore be useful to borrow information from other patients when data is lacking. Using a nonlinear mixed effect model (NLME) [17] framework, the growth and decay rates of all patients can be estimated together, allowing parameter estimates to be guided towards the population mean in the absence of clear information. Bayesian implementations of NLME models allow measuring the uncertainty of parameter estimates for each patient while also quantifying the spread of parameter values in the population. Moreover, Bayesian models can be readily expanded to look for correlations between patient-specific covariates and parameter values, all within the uncertainty quantifying framework [22].

In this work, we introduce a hierarchical Bayesian model of subpopulation dynamics that includes covariate effects from baseline and on-treatment covariates to two interpretable model parameters: the growth rate *ρ*^*r*^ and the initial proportion *π*^*r*^ of the resistant subpopulation. If any covariates are correlated with a higher *π*_*r*_ and *ρ*_*r*_, then modeling covariate effects should improve predictions of future M protein values compared to an NLME model without covariate effects. To avoid overfitting weak or spurious correlations, a sparsity-enforcing prior is used to penalize the covariate effects. More details about the hierarchical Bayesian model of subpopulation dynamics are provided in the Supplementary material.

Baseline and on-treatment covariates from each patient were pre-processed depending on the covariate type. Categorical covariates with L levels were turned into L-1 binary covariates (so-called one-hot encoding). Numerical covariates with continuous values like age and weight were categorized and one-hot encoded. The treatment group was included as a binary covariate called “Treat is Kd”, with a value of “1” indicating treatment with carfilzomib and dexamethasone. From numerical longitudinal measurements, a set of summarizing functions were used to extract covariates from each time series. The supplemental information contains more details, including a list of summary functions and longitudinal covariates.

### Model overview and comparison

Three models were compared by their ability to predict relapse. A hierarchical Bayesian model of subpopulation dynamics without covariate effects, a hierarchical Bayesian model of subpopulation dynamics with covariate effects, and a model based on linear extrapolation of M protein values inspired by [14] and [15]. This model, which we call the model of linear extrapolation, estimates the rate of change in M protein from the last two M protein measurements, and uses this to predict future M protein values. More details are provided in the supplementary information. By providing each model with M-protein measurements taken between the start of treatment and a chosen cutoff, the ability to predict relapse within six cycles following the cutoff was evaluated. Finally, relationships between model parameters and PFS were assessed and the importance of baseline and longitudinal covariates was evaluated and reported.

Figure 2 a) visualizes how the three models work, and Figure 2 b)-d) shows model predictions for three simulated example patients. The patients are highly heterogeneous in their treatment response and time to relapse. While patient 2 experiences a lasting response, patients 1 and 3 relapse during the observation period, with resistant populations growing at different rates. While the model of linear extrapolation is highly sensitive to observation noise, the hierarchical Bayesian models of subpopulation dynamics fit the observed data well while quantifying the uncertainty in model fit and prediction.

**Figure 2.**
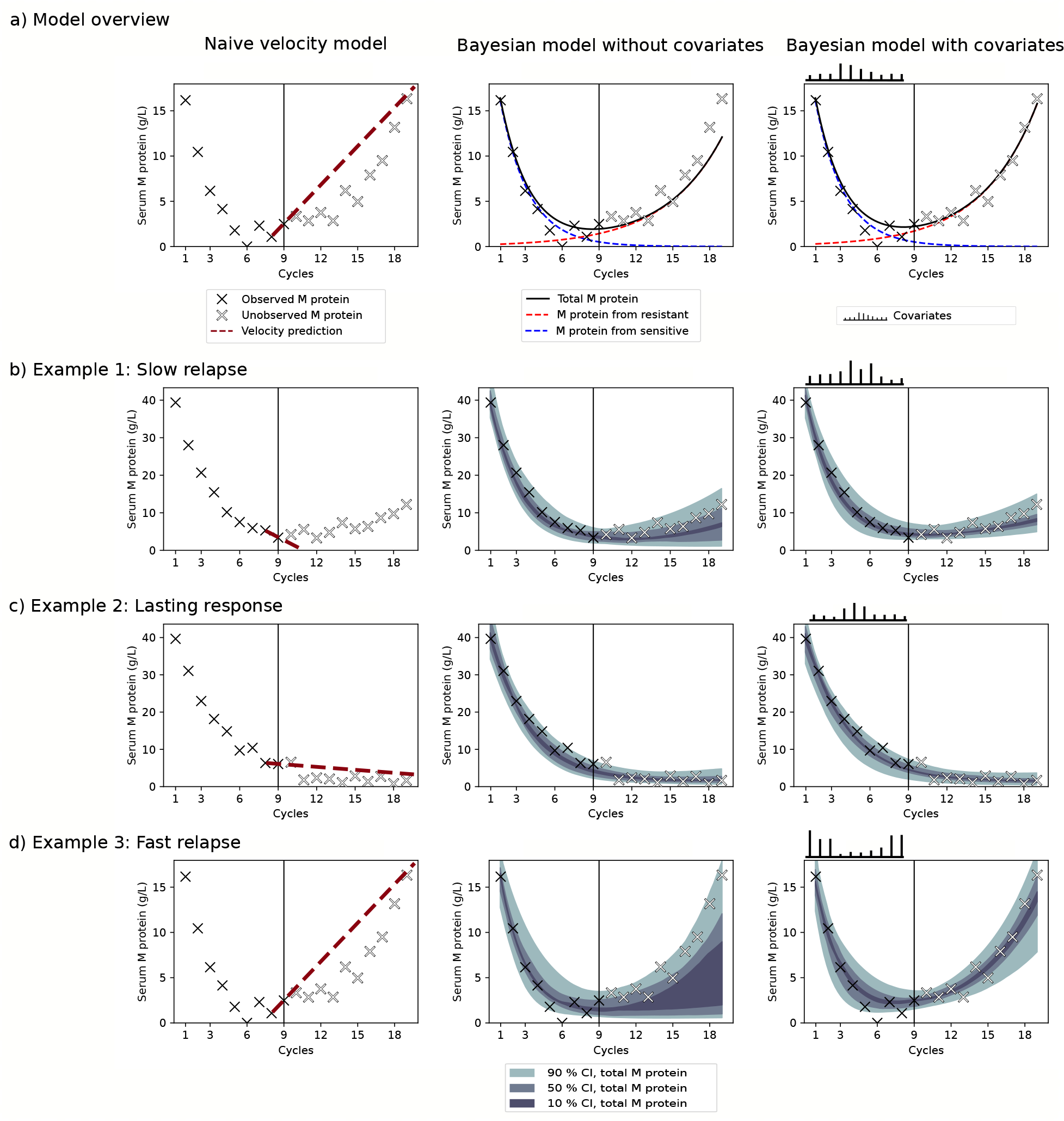
a) Overview of the three models fitted to observed M protein (black crosses) and used to predict unobserved M protein (white crosses). b)-d) simulated example patients with simulated covariate effects illustrate how each model fits to members of a heterogeneous population. Posterior credible intervals (CI) quantify the uncertainty of the hierarchical Bayesian model of subpopulation dynamics predictions. Both of the hierarchical Bayesian models of subpopulation dynamics predict future M protein measurements accurately for patients 1 and 2. Within the simulated population, patients with covariates similar to those of patient 3 have resistant subpopulations with higher growth rates. This is detected by the hierarchical Bayesian model of subpopulation dynamics with covariates, which predicts that patient 3 will experience a quick relapse.

### Statistical analysis

The computational intensity of model fitting, demanding several hours on four parallel central processing units (CPUs) for a single fit, hindered the full implementation of a bootstrap procedure with enough bootstrap samples to generate confidence intervals for the evaluation metrics. Instead, model performance was assessed based on six bootstrap samples, and the average score for each evaluation metric was reported. To generate a bootstrap sample from a population of *N* patients, *N* patients were sampled with replacements to create the patient set used for training, and the remaining out-of-bag [23] patients were used as test patients. The NLME framework was used to estimate the parameters of all patients together, using information from the entire population when estimating the parameters for each patient. For patients allocated to the training set, all M protein measurements were used. For patients in the test set, only M protein measurements taken during the chosen observation period were used to fit the model. After fitting the model to the data, the resulting posterior contained credible intervals for each population-level and individual parameter, and probabilistic forecasts of future M protein development for each patient.

All statistical analysis was performed in Python version 3.11.3. Posterior sampling was conducted in PyMC version 5.1.1 [24] using the No-U-Turn sampler (NUTS) introduced in [25]. To improve convergence, the sampler was initialized using automatic differentiation variational inference (ADVI) [26] within the PyMC software. To evaluate differences in average posterior median model parameters *ρ*^*r*^ and *π*^*r*^ between different levels of baseline covariates, the false discovery rate was controlled at 0.25 using the Benjamini-Hochberg method.

### Code and data availability

The code for this project is available at https://github.com/evenmm/mm-predict-ikema. Data from the IKEMA trial (NCT03275285) can be requested through the data-sharing platform Vivli.

## Results

### Prediction of relapse within six cycles

The ability to predict relapse in M protein within the next six cycles was evaluated for the model of linear extrapolation, the hierarchical Bayesian model of subpopulation dynamics without covariate effects, and the hierarchical Bayesian model of subpopulation dynamics with covariate effects. The following subset of the International Myeloma Working Group (IMWG) criteria [27] for progressive disease was used as relapse criteria. Any one or more of the following criteria: a) Increase of 25% from lowest confirmed response value in Serum M-protein (absolute increase must be ≥ 0.5 g/dL); b) Serum M-protein increase ≥ 1 g/dL, if the lowest M component was ≥ 5 g/dL. Since the model only relates to serum M protein, the definition of relapse used in this evaluation excludes relapse from causes other than M protein. Figure 3 a) visualizes the relapse criteria together with observed M protein and predicted credible interval for a simulated example patient.

**Figure 3.**
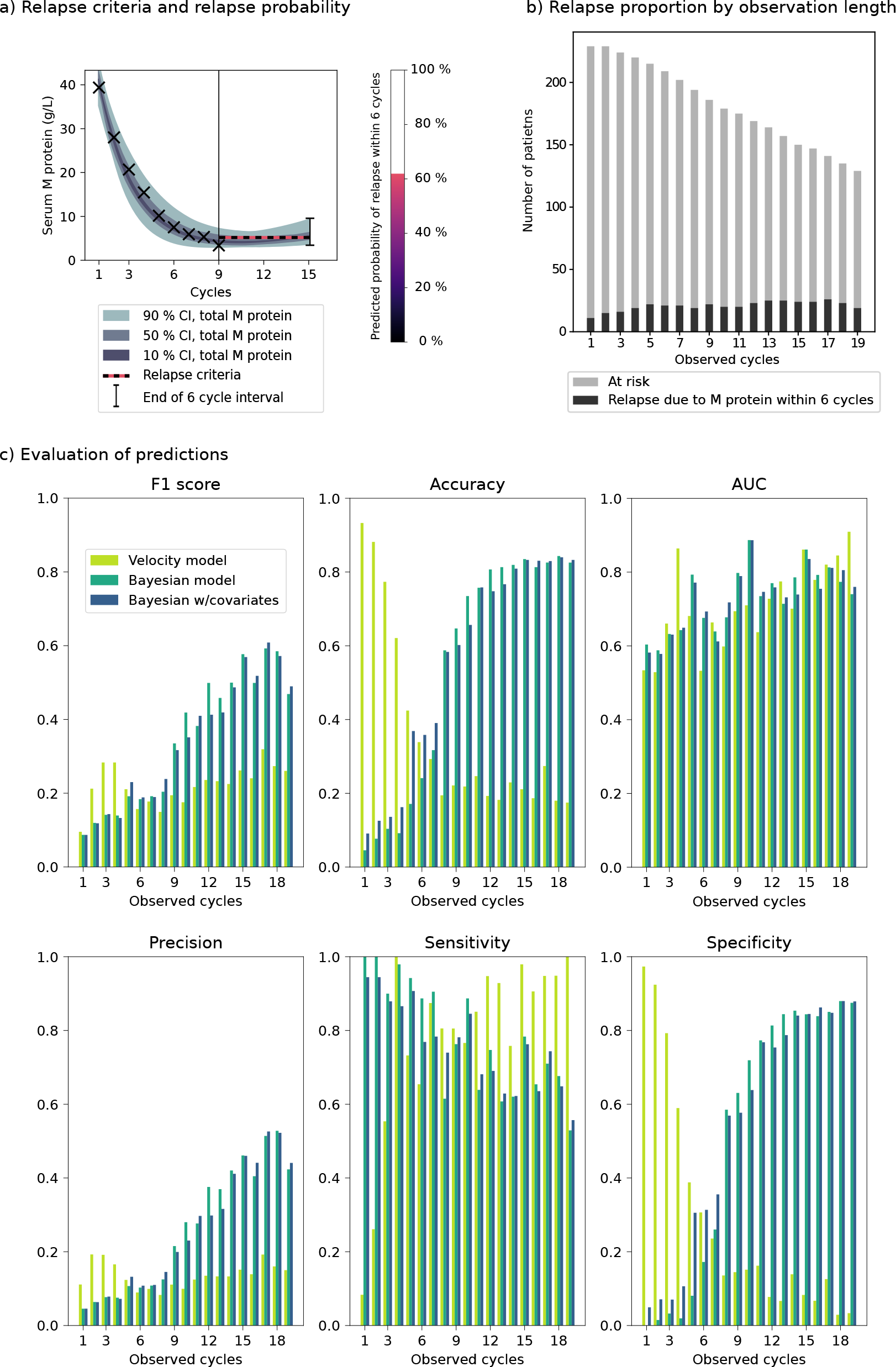
a) Illustration of relapse criteria and relapse probability. b) Population size and number of patients about to relapse by the number of observed cycles. c) Average area under the curve (AUC), accuracy, F1 score, precision, sensitivity, and specificity across six bootstrap samples for observation period lengths ranging from 1 to 19 cycles.

Since patients who relapse, are censored, or die during the observed period must be removed from the population, the number of patients at risk decreases with increasing length of the observation period. The number of patients who relapse within the next six cycles also varies with the length of the observation period. Figure 3 b) shows the decrease in the study population and variation in relapse proportion across different observation period lengths.

In Figure 3 a), the predicted probability of relapse equates to the proportion of the credible interval that lies above the relapse criteria after six cycles. To classify whether a patient will relapse or not within the next six cycles, a probability threshold is needed to turn the probability into either a “1” for relapse or “0” for no relapse. For each model, a single threshold was chosen and used for all observation window lengths. Figure 3 c) shows the average prediction performance across six bootstrap samples for observation windows of different lengths. Model comparisons under two alternative choices of threshold are shown in the supplementary materials.

### Model parameters predict PFS

A survival analysis showed that the estimated model parameters for the growth rate of the resistant subpopulation and proportion of resistant cells at the start of treatment separates patients into groups with highly different PFS. Figure 4 a)-c) shows Kaplan-Meier plots for this analysis. Cox regressions showed that patients with growth rate *ρ*^*r*^ of the resistant subpopulation above the median had a shorter PFS compared to below median (*p <* 2 10^−16^) and that patients with initial proportion *π*^*r*^ of the resistant subpopulation above the median had a shorter PFS compared to below median (*p* = 2 10^−13^). Notably, no difference in PFS between high and low values of *ρ*^*s*^ was found (*p* = 0.8). For this analysis, the PFS definition was the same as in the IKEMA trial, which included relapse from causes other than serum M protein.

**Figure 4.**
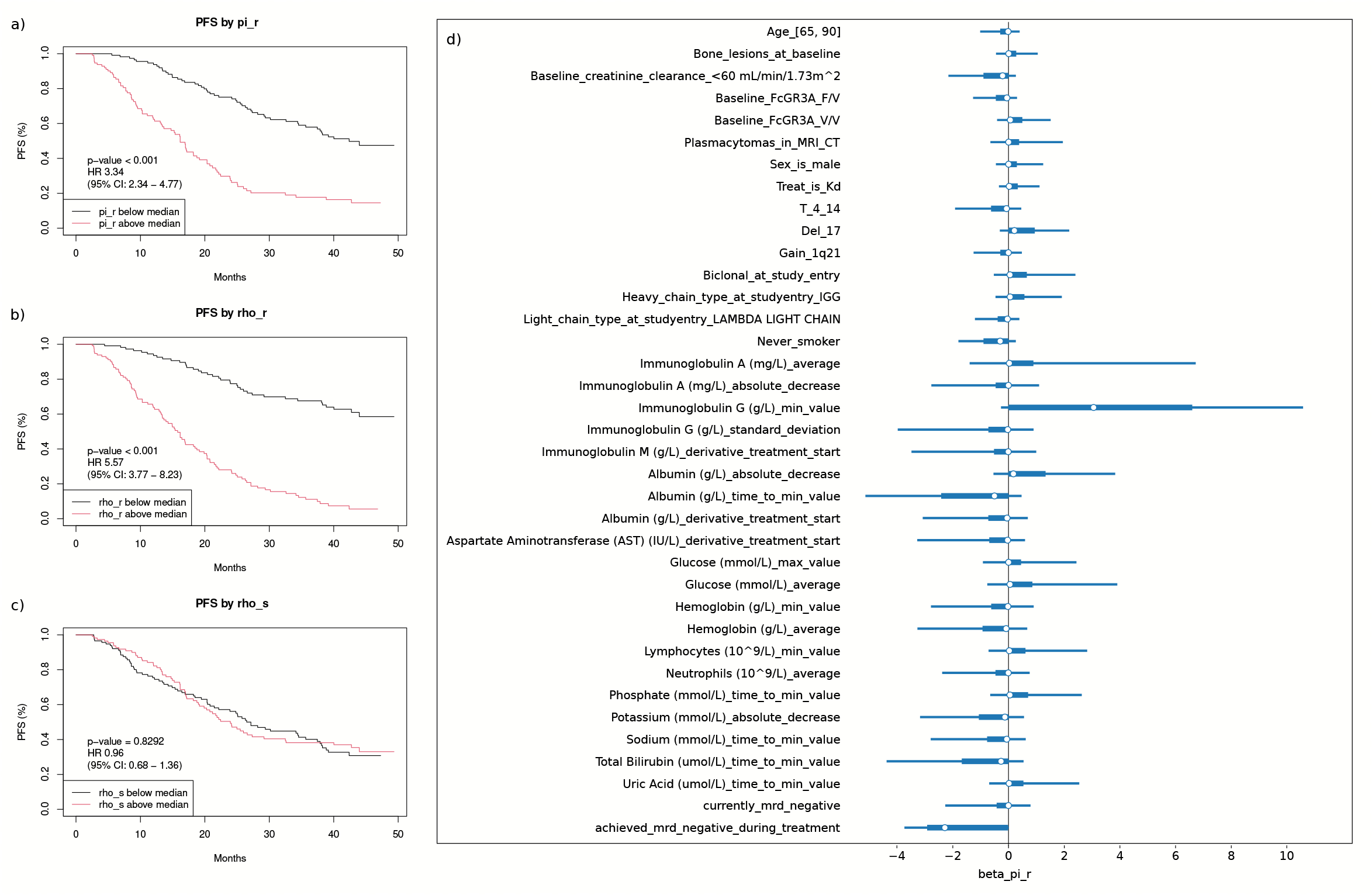
a-c) Kaplan Meier plot of PFS, by high and low values of *π*^*r*^, *ρ*^*r*^, and *ρ*^*s*^, separately. d) 95 % posterior credible intervals for coefficients between selected covariates and initial proportion of resistant subpopulation *π*^*r*^.

### Importance of baseline and longitudinal covariates

In the hierarchical Bayesian model of subpopulation dynamics with covariate effects, the covariate ‘achieved MRD negative during treatment’ was associated with a smaller estimated proportion of resistant cells at the start of treatment *π*^*r*^. Additionally, patients with larger minimum values of Immunoglobulin G during observation tended to have larger estimated *π*^*r*^. Figure 4 d) shows 95 % credible intervals for a selection of covariate effects on *π*^*r*^. All covariate effects on *π*^*r*^ and *ρ*^*r*^ are shown in supplementary figures S1 and S2.

Separately, a comparison of average posterior median model parameters *π*^*r*^, *ρ*^*r*^, and *ρ*^*s*^ found that patients with plasmacytomas in MRI or CT at baseline on average had a significantly higher estimated *π*^*r*^ than patients without. This was the only significant difference when correcting for multiple testing. The difference in average posterior median *π*^*r*^ is shown in Table 1. None of the baseline covariates had a significant effect on posterior median *ρ*^*r*^ or *ρ*^*s*^.

**Table 1:**
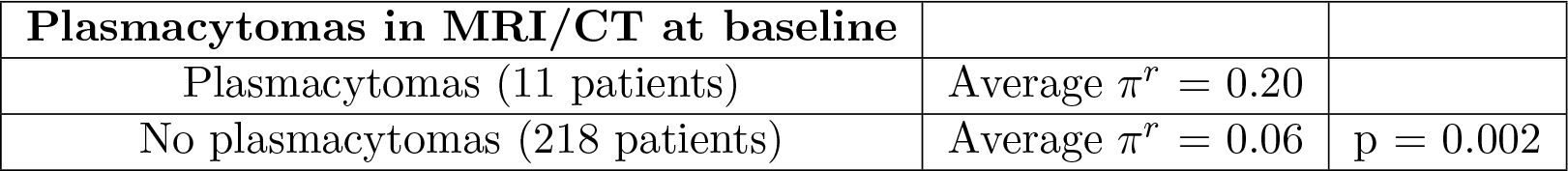
Significant differences in average posterior median *π*^*r*^.

## Discussion

Evaluation of relapse prediction for the next six cycles in Figure 3 c) showed that after observing at least eight treatment cycles, the hierarchical Bayesian models of subpopulation dynamics predict relapse within six cycles with an average sensitivity between 60 and 80 % and average specificity between 70 and 80 %. After observing at least nine treatment cycles, the Bayesian mechanistic model achieved AUC scores ranging from 0.71 to 0.89. This matches or exceeds previous work [14] on relapse prediction in patients treated with combinations of bortezomib, lenalidomide, and dexamethasone, which achieved an AUC score of 0.83.

In the F1 score, which is the harmonic mean of precision and sensitivity, the hierarchical Bayesian models of subpopulation dynamics performed better than the model of linear extrapolation after observing at least six cycles. Three aspects of the hierarchical Bayesian model of subpopulation dynamics model may explain why it outperforms the model of linear extrapolation. Firstly, the model of linear extrapolation is unable to predict an increasing growth rate in M protein, which is common in relapsing patients. In contrast, the hierarchical Bayesian model of subpopulation dynamics encodes this behavior explicitly through the mathematical model. Secondly, the inherent handling of uncertainty makes the hierarchical Bayesian model of subpopulation dynamics robust to noisy observations. Finally, the statistical framework lets the hierarchical Bayesian model of subpopulation dynamics learn trends from patients in the training set. How well this performance generalizes to patients with different treatment histories undergoing different treatment regimens requires further evaluation in another cohort. If the performance is found to generalize to other patient cohorts, our model could be used to guide the frequency of follow-up measurements and to alert clinicians ahead of time when patients are about to relapse.

The model of linear extrapolation achieves very high accuracy and specificity scores for 1 to 4 cycles long observation windows. This is explained by the fact that most patients have decreasing M protein values at the start of treatment. Therefore, even in the presence of observation noise, cases where M protein increases are very unlikely unless the disease is progressing. As more data are observed, the accuracy and specificity of the model of linear extrapolation drops. For longer observation windows, M protein values tend to be lower and more stable, meaning that measurement error may randomly make the M protein appear to be increasing. This explains the observed drop in specificity and accuracy. The initially very low sensitivity is explained by the inability of the model of linear extrapolation to predict relapse in patients with decreasing M protein values. The accuracy and specificity of the hierarchical Bayesian model of subpopulation dynamics increases with the length of the observation period, accompanied by a modest decrease in sensitivity. Consequently, the model of linear extrapolation is recommended when less than six cycles are observed, and the hierarchical Bayesian model of subpopulation dynamics is recommended for longer observation periods.

As seen in Figure 3 b), there are fewer patients at risk after longer observation periods. Since the model is trained independently for each observation period length, fewer patients should lead to a reduction in model accuracy. However, longer observation times mean more M protein measurements per patient, and the population becomes more homogeneous by selecting patients who have survived without relapse. Figure 3 shows that the scores of the hierarchical Bayesian model of subpopulation dynamics are very low with few observations, yet increase with the length of the observation windows, outperforming the model of linear extrapolation. From this we can conclude that having more M protein measurements and a more homogeneous population makes up for the loss in population size.

Since the hierarchical Bayesian models of subpopulation dynamics with and without covariate effects had comparable predictive power across different observation period lengths, we conclude that incorporating covariate effects did not improve the predictive capacity of the hierarchical Bayesian model of subpopulation dynamics. This means that none of the baseline covariates included in this work were informative about individual M protein trajectories. This does not, however, exclude the possibility that other covariates measured with novel or more detailed sequencing techniques could hold important information and thus predictive value.

The survival analysis in Figure 4 shows that separating patients by high and low values of the mechanistic parameters *ρ*^*r*^ (growth rate of resistant population) and *π*^*r*^ (initial fraction of resistant population) differentiates PFS. For this analysis, the PFS definition was not limited to relapse from M protein but included death and relapse from other causes. This shows that the estimated model parameters describing M protein development hold important information about the patient’s status. Moreover, this is true even when including patients that relapse from other causes or die before progressing due to M protein.

While including covariate information did not improve predictive accuracy, patients who achieved MRD negativity during observation tended to have a smaller proportion of resistant cells at the start of treatment. As MRD is arguably the strongest marker of long-term survival outcome in MM [28], it is reassuring to find that our model estimates a lower proportion of resistant cells at the start of treatment for patients who achieve MRD negativity during observation. The results of the covariate effect analysis should be viewed as exploratory. The difference in *π*^*r*^ between patients with or without Plasmacytomas in MRI or CT at baseline was not captured by our model. None of the covariate effects captured by our model were found to be significant in the separate comparison of average model parameters when correcting for multiple testing.

The model assumes that a resistant subpopulation is present from the start of treatment.

Two possible alternative explanations are an outgrowth of resistant clones during treatment, and phenotypic switching caused by cell plasticity [29, 30]. In practice, should resistant clones arise during treatment, the hierarchical Bayesian model of subpopulation dynamics should be able to fit such a patient by estimating an initial resistant population of negligible size. Consequently, our model is not able to distinguish between these two cases, nor should this alternative be detrimental to the model’s performance.

As a necessity of the 28-day interval between M protein measurements, M protein and population sizes are modeled as smoothly changing values, despite treatment being given intermittently at each 28-day cycle. Mechanics of differential clearance and M protein decay rates are also excluded from the model. In the time between M protein measurements, there may be time for cells to switch between phenotypes, by intermittent dormancy at treatment administrations followed by growth between doses. Nevertheless, initial decay in M protein followed by an increase is indicative of a change in the clonal composition of the cancer. Therefore, the model may prove capable of response prediction regardless of the actual mechanisms at play.

In this study, a single threshold used for all observation window lengths was chosen for each model. Since the population at risk changes with the length of the observation window, the optimal threshold value may vary with the length of the observation period. The supplementary material provides a model comparison where the probability thresholds were set either to optimize the F1 score or to ensure a sensitivity of 80 %. This lets the threshold vary with the length of the observation period but does not provide a representative view of the model performance, since the labels in the test set are used to set the threshold in each case. Provided with a larger dataset, a validation set could have been used to set the threshold value instead, which would give an unbiased estimation of the performance on the test set while allowing different thresholds for different observation lengths.

In this work, a mathematical model has been developed that predicts relapse with acceptable sensitivity and specificity after observing eight or more treatment cycles. This demonstrates the usefulness of mathematical models for predicting clinical events. The generalizability of the prediction accuracy to other regimens and/or patients with different inclusion criteria requires evaluation in other cohorts.

## Data Availability

The code for this project is available at https://github.com/evenmm/mm-predict-ikema. Data from the IKEMA trial (NCT03275285) can be requested through the data-sharing platform
Vivli.

https://search.vivli.org/studyDetails/fromSearch/42d32e57-51ce-4d54-9613-eb3f7c73d30e

https://github.com/evenmm/mm-predict-ikema

## Acknowledgments

This publication is based on research using data from data contributor Sanofi that has been made available through Vivli, Inc. Vivli has not contributed to or approved, and is not in any way responsible for, the contents of this publication. Simulation studies were performed on resources provided by UNINETT Sigma2 - the National Infrastructure for High-Performance Computing and Data Storage in Norway, and by Oslo Centre for Biostatistics and Epidemiology. The study has benefitted from data provided by Oslo Myeloma Center and the COMMPASS project to sharpen questions and develop models. We thank Anna Lysén at Oslo Myeloma Center for her aid in providing access to data which inspired the development of the models presented in this work. E. M. Myklebust, A. Köhn-Luque, and A. Frigessi were supported by the Center for research-based-innovation BigInsight under grant 237718 by the Research Council of Norway. J. Foo and K. Leder were supported by the Fulbright US-Norway Foundation. J. Foo and K. Leder were supported by the University of Oslo-University of Minnesota Norwegian Centennial Chair Grant. J. Foo was supported by the US National Science Foundation under grant number DMS-2052465. K. Leder was supported by the US National Science Foundation under grant number CMMI-2228034. We acknowledge funding from the Research Council of Norway through projects DL: Pipeline for individually tailoring new treatments in hematological cancers (PINpOINT) under project number 294916; INTPART-International Partnerships for Excellent Education and Research under project number 309273; and Integreat - Norwegian Centre for knowledge-driven machine learning under project number 332645. The authors also acknowledge the Centre for Digital Life Norway for supporting the partner project PINpOINT.

## CRediT author contribution statement

**E. M. Myklebust:** Conceptualization, Methodology, Software, Validation, Formal analysis, Investigation, Data curation, Writing – original draft, Writing – review & editing, Visualization. **F. Schjesvold:** Conceptualization, Methodology, Writing – review & editing. **A. Frigessi:** Conceptualization, Methodology, Writing – review & editing, Supervision, Funding acquisition. **K. Leder:** Conceptualization, Methodology, Writing – review & editing, Supervision. **J. Foo:** Conceptualization, Methodology, Writing – review & editing, Supervision. **A. Köhn-Luque:** Conceptualization, Methodology, Writing – original draft, Writing – review & editing, Supervision.

