## Supplementary information for "Relapse prediction in multiple myeloma patients treated with isatuximab, carfilzomib, and dexamethasone"

#### 1 Evaluation of predictions with different thresholds

The choice of probability threshold impacts the predictive performance of the model. In the main article, the thresholds were set to 0.02 for the NLME models with and without covariates and 0.45 for the model of linear extrapolation. Figure 1 shows the average performance across 6 bootstrap samples under two alternative choices of probability threshold. In both cases, the thresholds were set using information from the test set. Therefore, it does not represent an unbiased view of the predictive performance but can be used to compare the performance of the methods. The AUC score does not depend on a threshold. Therefore it remains the same and is not repeated here.

In Figure 1 a), for each number of observed cycles, the threshold was chosen to achieve 80 % sensitivity on average, reflecting a prioritization of relapse discovery at a potential cost of specificity. With this choice of threshold, the models can be compared by their compared by the cost in specificity incurred by requiring a sensitivity of 80 %. Both hierarchical Bayesian models of subpopulation dynamicswith and without covariates have higher specificity than the model of linear extrapolation for 14 out of 19 observation window lengths. The NLME models with and without covariates are comparable across all metrics and observation period lengths.

Alternatively, the probability threshold that maximizes the F1 score can be chosen. Figure 1 b) shows the average performance across 6 bootstrap samples with this threshold. The F1 scores tend to increase with the number of observed cycles for all three models. Both hierarchical Bayesian models of subpopulation dynamicshave higher F1 score than the model of linear extrapolation for 13 out of 19 observation window lengths. The F1 score of the NLME models with and without covariates are comparable across all observation period lengths.

#### 2 Baseline and longitudinal covariates

Table 1 lists the baseline covariates. Table 2 lists the summary functions used to extract summary covariates from longitudinal measurements. Table 3 lists the longitudinal measurements

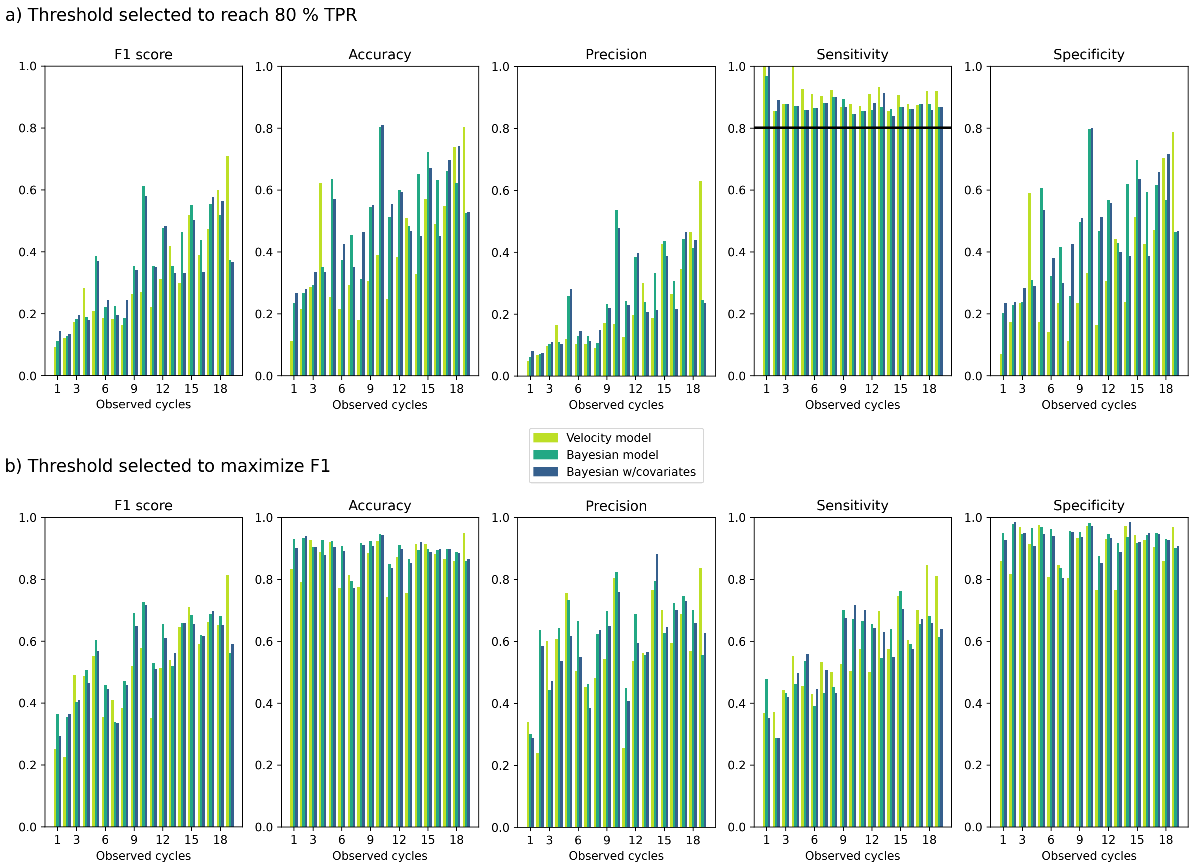


Figure 1: Evaluation metric scores with two alternative ways to set the probability threshold to classify whether patients will relapse or not. In a), the smallest threshold that achieved an average sensitivity of 80 % across all folds is used, for each observation window length separately. In b), the threshold that maximizes the F1 score is chosen for each observation window length separately.

from which summary covariates were extracted. Figures 2 and 3 show all covariate effects on $\pi^r$ and $\rho^r$.

Table 1: List of baseline covariates.

| |
|---|
| Age |
| Race |
| Sex |
| Region |
| Weight |
| ISS stage at study entry |
| Duration of last treatment |
| Number of prior lines of treatment |
| Treatment group (isatuximab or control) |
| T_4_14 |
| T_14_16 |
| Del 17 |
| Gain 1q21 |
| Chromosomal aberration risk |
| Biclonal at study entry |
| Heavy chain type at study entry |
| Light chain type at study entry |
| Received radiotherapy |
| Time from diagnosis to randomization |
| Presence of bone lesions at baseline |
| Baseline Serum M protein |
| Baseline Urine M protein |
| Baseline plasma cell bonemarrow |
| Baseline creatinine clearance |
| Baseline ECOG status |
| Baseline FcGR3A |
| Plasmacytomas in MRI or CT at baseline |
| Baseline LDH |
| Baseline smoke status |

Table 2: List of summary functions for extracting covariates from longitudinal measurements.

| | |
|---|---|
| minimum value | min(x). |
| Maximum value | max(x). |
| Average | sum(x)/len(x). |
| Standard deviation | $\sqrt{\frac{1}{len(x)}\text{sum}_{i=1}^{len(x)}(x_i - mean(x))^2}$. |
| Absolute decrease | x[0] - min(x). |
| Time to minimum value | Time from start of treatment to observing min(x). |
| Derivative at start of treatment | $\frac{x[1]-x[0]}{t[1]-t[0]}$ |
| Derivative at end of observation period | $\frac{x[-1]-x[-2]}{t[-1]-t[-2]}$ |

Table 3: List of longitudinal covariates, with units.

| | |
|---|---|
| Immunoglobulin A | mg/L |
| Immunoglobulin G | mg/L |
| Immunoglobulin M | mg/L |
| Kappa light chain, Free/Lambda light chain, free | Ratio |
| Kappa light chain, Free | mg/L |
| Lambda light chain, free | mg/L |
| Alanine Aminotransferase | IU/L |
| Albumin | g/L |
| Alkaline Phosphatase | IU/L |
| Aspartate Aminotransferase | IU/L |
| Bicarbonate | mmol/L |
| Calcium | mmol/L |
| Chloride | mmol/L |
| Creatinine | $\mu$mol/L |
| Direct Bilirubin | $\mu$mol/L |
| GFR from Creatinine adjusted for BSA | mL/min/1.37$m^2$ |
| Glucose | mmol/L |
| Hemoglobin | g/L |
| Leukocyte count (WBC) | $10^9/L$ |
| Lymphocytes | $10^9/L$ |
| Magnesium | mmol/L |
| Neutrophils | $10^9/L$ |
| Phosphate | mmol/L |
| Platelet count | $10^9/L$ |
| Potassium | mmol/L |
| Sodium | mmol/L |
| Total Bilirubin | $\mu$mol/L |
| Urea Nitrogen (BUN) | mmol/L |
| Uric acid | $\mu$mol/L |

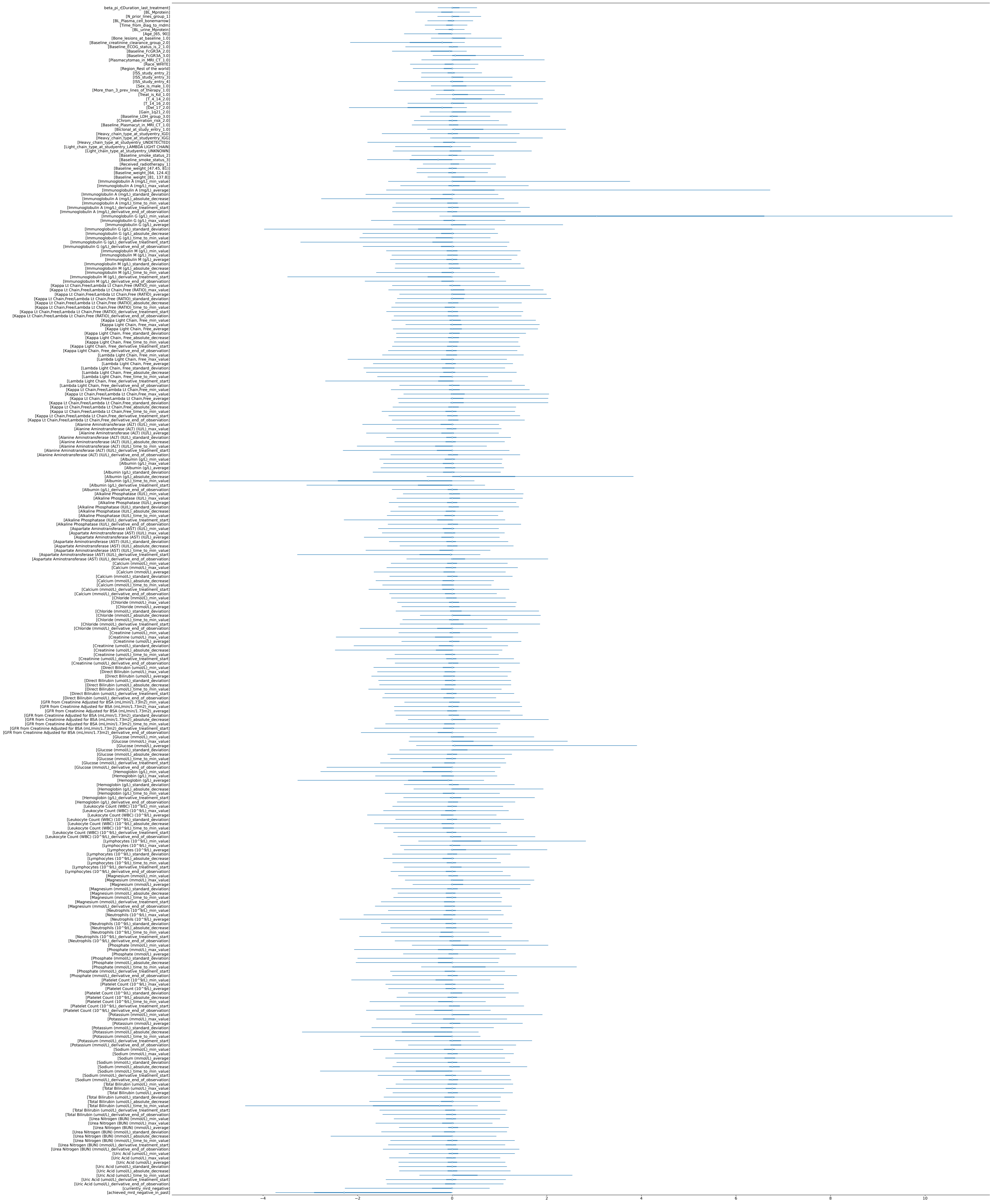

Figure 2: All covariate effects on $\pi^r$.

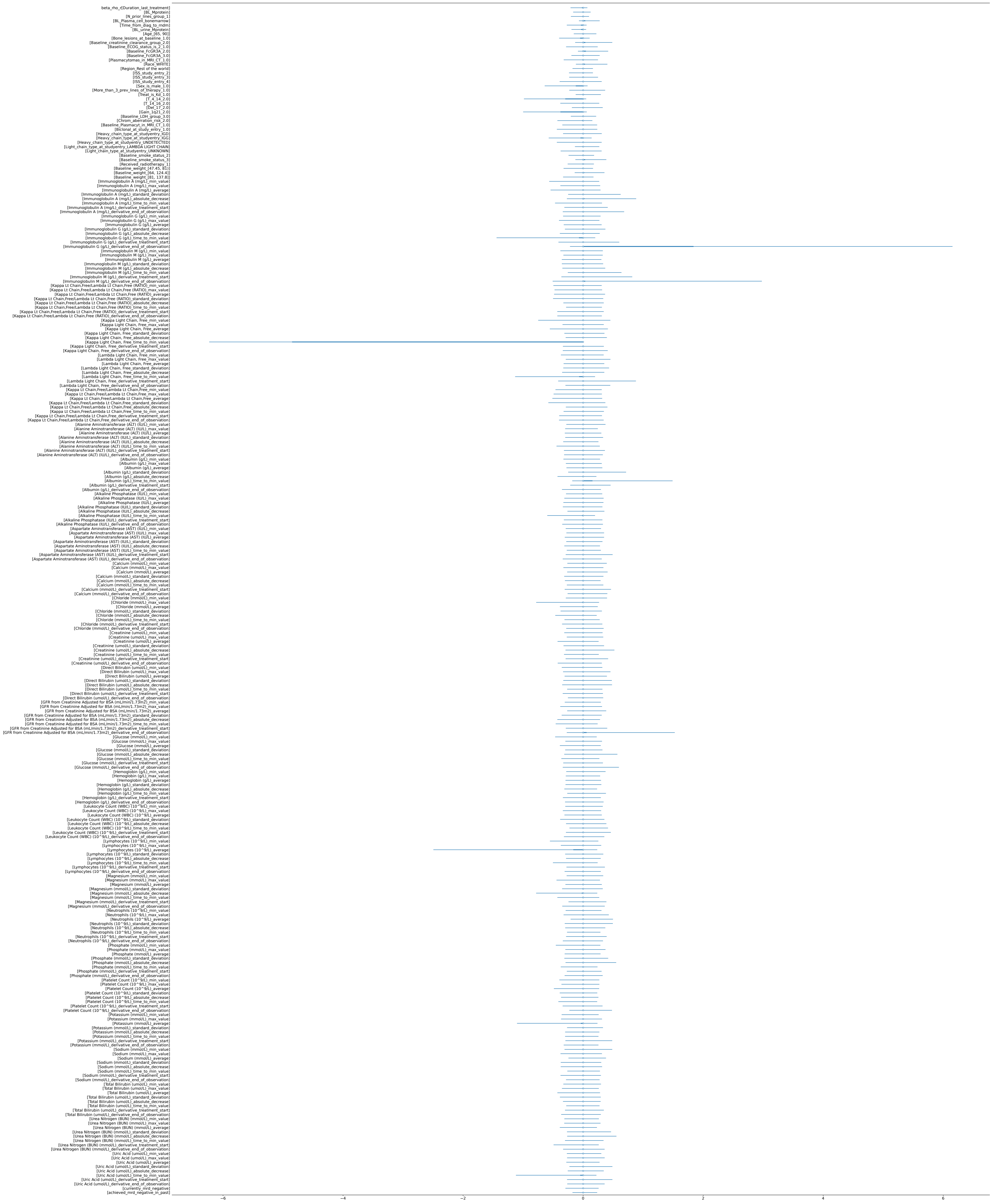


Figure 3: All covariate effects on $\rho^r$.

### 3 Model of linear extrapolation

As a baseline method, the model of linear extrapolation predicts future M protein values by linear extrapolation using the last two observed M protein measurements. To account for measurement error, 100 perturbed observations are created for each of the two points separately by adding noise terms sampled from a normal distribution with zero mean and standard deviation 0.5 g/dL, which equals the minimum change required for progressive disease. Then, 100 different velocities are calculated from the perturbed points and used to make a simple forecast of the tumor progression.

### 4 Mathematical model of treatment response and relapse

With $N$ as the total number of patients, assign to each patient an index $i \in [1, \ldots, N]$, and denote their $M_i$ measured M protein values as $y_i = (y_{i1}, \ldots, y_{iM_i}) \in \mathbb{R}^{M_i}$, where each measurement $y_{ij}$ is taken at the corresponding time $t_{ij}$ in $t_i = (t_{i1} = 0, \ldots, t_{iM_i}) \in \mathbb{R}^{M_i}$. Additionally, let the $p$ covariates measured at baseline be denoted as $x_i = (x_{i1}, \ldots, x_{ip}) \in \mathbb{R}^p$. The observed M protein $y_i$ is modeled as the sum of a resistant population growing exponentially with rate $\rho_i^r$ and a sensitive population decaying exponentially with rate $\rho_i^s$, with initial total population size $M_i^0$ and initial proportion of resistant cells $\pi_i^r$. Observation noise terms $\varepsilon_{ij}$ are assumed to be normally distributed with variance $\sigma^2$.

$$\begin{aligned} y_{ij} &= M(t_{ij}, \theta^i) + \varepsilon_{ij} \ , \ \ \varepsilon_{ij} \sim \mathcal{N}(0, \sigma^2) \\ M(t_{ij}, \theta^i) &= M_i^0 \pi_i^r \exp\left(\rho_i^r t_{ij}\right) + M_i^0 (1 - \pi_i^r) \exp\left(\rho_i^s t_{ij}\right) \end{aligned} \tag{1}$$

Notice that since $t_{i1} = 0$, $y_{i1} = M(0, \theta_i) + \varepsilon_{i1} = M_i^0 + \varepsilon_{i1}$. The patient-specific mechanistic parameters in this model are:

- $M_i^0$: True M-protein value at the time when treatment is started.
- $\pi_i^r$: Fraction of resistant cells at the time when treatment is started.
- $\rho_i^r$: Growth rate of resistant cells under current treatment.
- $\rho_i^s$: Growth rate of sensitive cells under the current treatment.

Due to the physical constraints on the mechanistic parameters $\rho_i^s, \rho_i^r$, and $\pi_i^r$, we use a variable transform to work with model parameters that have support on the entire real line.

$$\text{Mechanistic parameters} \begin{bmatrix} \rho_i^s \\ \rho_i^r \\ \pi_i^r \end{bmatrix} = \begin{bmatrix} -\exp(\theta_{1i}) \\ \exp(\theta_{2i}) \\ \frac{1}{1+e^{-\theta_{3i}}} \end{bmatrix} \text{with domains} \begin{matrix} [-\infty, 0] \\ [0, \infty] \\ [0, 1] \end{matrix} .$$

$$\text{Model parameters } \theta^i = \begin{bmatrix} \theta_{1i} \\ \theta_{2i} \\ \theta_{3i} \end{bmatrix} = \begin{bmatrix} \log(-\rho_i^s) \\ \log(\rho_i^r) \\ \log(\frac{\pi_i^r}{1-\pi_i^r}) \end{bmatrix} \in \mathbb{R}^3.$$

#### 4.1 Nonlinear mixed effect model (NLME)

A nonlinear mixed effect (NLME) framework was used to estimate the parameters of all patients together, using information from the entire population when estimating the parameters for each patient. For an introduction to Bayesian nonlinear mixed effects models for repeated measurements, see [1]. After fitting the model to the data, the resulting posterior contains credible intervals for each population-level and individual parameter and probabilistic forecasts of future M protein development for each patient. In the NLME model, individual parameters $\theta_{1i}$, $\theta_{2i}$, and $\theta_{3i}$ are modeled as normally distributed around population means $\alpha_1$, $\alpha_2$, and $\alpha_3$, with patient-specific random intercepts allowing the patient-specific $\rho_i^s, \rho_i^r$, and $\pi_i^r$ to deviate from the group means to match the observed M protein. This works as a prior that encourages parameters to be more similar to the group averages in the absence of M protein observations that indicate otherwise.

$$\text{For } l = 1, \dots, 3: \quad \theta_{li} \sim \mathcal{N}(\alpha_l, \omega_l^2) \tag{2}$$

We do not seek to predict $M_i^0$ using the covariates since we believe there is enough information in the observed M protein alone to estimate it. To deal with the requirement that all M protein values be greater than or equal to zero, we assign a prior to the log-transformed $\theta_{4i} = \log(M_i^0)$:

$$\begin{aligned} \theta_{4i} &\sim \mathcal{N}\left(\log(y_{i1}), \xi^2\right) \\ M_i^0 &= \exp(\theta_{4i}) \end{aligned} \tag{3}$$

The parameters $\alpha_l$, $l = 1, 2, 3$ are given normal priors centered around suitable guesses $\widehat{\alpha_l}$ of the average $\theta_{li}$ in the population, for $l = 1, 2, 3$. The variance of this prior is set to 1 to cover a wide enough range of values.

$$\alpha_l \sim \mathcal{N}(\widehat{\alpha_l}, 1) \ , \ l \in [1, 2, 3] \tag{4}$$

We chose non-informative priors restricted to positive values for the standard deviations $\sigma$ and $\omega$.

$$\begin{aligned} \sigma &\sim \text{HalfNormal}(0,1) &: \ & \sigma > 0 \\ \xi &\sim \text{HalfNormal}(0,1) &: \ & \xi > 0 \\ \omega_l &\sim \text{HalfNormal}(0,1) &: \ & \omega_l > 0 \ , \ l \in [1, 2, 3] \end{aligned} \tag{5}$$

##### 4.2 Incorporating covariate information: NLME with linear covariate effects

In addition to the M protein data that the NLME models, every patient has a set of covariates that are measured at baseline or longitudinally. It is natural to ask what additional stratification these covariates can achieve beyond the information available through the M protein measurements. To answer this question, the NLME can be extended by adding one simple hierarchical layer. The purpose of this section is to describe what this layer is and how it can be used to include any stratifying covariates.

Let the covariate vector be denoted as $x$. In the linear covariate model, the effect of the $p$ covariates in $x$ on parameter $\theta_l$ is modeled by the function $f_l(x) = x\beta_l^T$. Note that $f_l(x)$ can be freely exchanged to other, more flexible mappings, each of which would complicate the convergence of the Bayesian inference in their own way. The parameters related to the speed of relapse are $\rho^r$ and $\pi^r$, so for the cause of simplicity, the model is extended with linear covariate effects for these two but not for $\rho^s$.

$$\text{For } l \in \{2, 3\}: \quad \theta_{li} \sim \mathcal{N}(\alpha_l + x_i\beta_l^T, \omega_l^2), \ \ \beta_l = (\beta_{l1}, \dots, \beta_{lp}) \in \mathbb{R}^p \tag{6}$$

Note that the inclusion of baseline covariate effects alters the interpretation of the $\alpha$ parameters, which do no longer represent population means, but must be interpreted as the expected parameter for a patient with all covariates equal to 0.

To include covariates in $x$ without knowing a priori how relevant they are to the parameters $\rho_i^s, \rho_i^r$, and $\pi_i^r$, we need a sparsity inducing prior on the coefficients $\beta_l$. In their 2010 paper, Carvalho et al.[2] introduced the Horseshoe prior, a regularization method that shrinks some parameters to zero while simultaneously allowing nonzero parameters by using heavy-tailed distributions, an approach that offers good computational tractability compared to the spike-and-slab prior. Piironen and Vehtari expanded upon this in 2017[3] by suggesting a hierarchical regularized horseshoe, presenting a complete hierarchical framework with stronger shrinkage for weakly identified parameters.

We use the hierarchical regularized horseshoe as a sparsity-inducing prior for the parameters $\beta_l$. This introduces a global shrinkage parameter $\tau$ plus one local shrinkage parameter $\tilde{\lambda}_{lb}$ for each $\beta_{lb}, b \in [1, \ldots, p]$, and requires an estimate of the number of nonzero parameters, $p_0$, which we set to $\widehat{p_0} = \text{int}(p/2)$. In addition, for sampling efficiency, we reparametrize $\beta_{lb}$ as $\beta_{lb} = z_{lb}\tau\tilde{\lambda}_{lb}$ to sample $z_{lb} \sim \mathcal{N}(0, 1)$ from a standard normal distribution.

$$\begin{aligned}
\beta_{lb} &\sim \mathcal{N}(0, \tau^2 \cdot \tilde{\lambda}_{lb}^2) \;\;,\;\; l \in [1, 2, 3] \;\;,\;\; b \in [1, \ldots, p] \\
\tau &\sim \text{Half-StudentT}_2 \left( \frac{p_0}{p - p_0} \cdot \frac{\sigma}{\sqrt{N}} \right) \;\;,\;\; l \in [1, 2, 3] \;\;,\;\; b \in [1, \ldots, p] \\
\tilde{\lambda}_{lb}^2 &= \frac{c^2 \lambda_{lb^2}}{c^2 + \tau^2 \lambda_{lb}^2} \\
\lambda_{lb} &\sim \text{Half-StudentT}_2(1) \;\;,\;\; l \in [1, 2, 3] \;\;,\;\; b \in [1, \ldots, p] \\
c^2 &\sim \text{InverseGamma}(1, 1)
\end{aligned} \tag{7}$$
